# Ancestry May Confound Genetic Machine Learning: Candidate-Gene Prediction of Opioid Use Disorder as an Example

**DOI:** 10.1101/2020.09.12.20193342

**Authors:** Alexander S. Hatoum, Frank R. Wendt, Marco Galimberti, Renato Polimanti, Benjamin Neale, Henry R. Kranzler, Joel Gelernter, Howard J. Edenberg, Arpana Agrawal

## Abstract

**Background:** Machine learning (ML) models are beginning to proliferate in psychiatry, however machine learning models in psychiatric genetics have not always accounted for ancestry. Using an empirical example of a proposed genetic test for OUD, and exploring a similar test for tobacco dependence and a simulated binary phenotype, we show that genetic prediction using ML is vulnerable to ancestral confounding.

**Methods:** We utilize five ML algorithms trained with 16 brain reward-derived “candidate” SNPs proposed for commercial use and examine their ability to predict OUD vs. ancestry in an out-of-sample test set (N=1000, stratified into equal groups of n=250 cases and controls each of European and African ancestry). We rerun analyses with 8 random sets of allele-frequency matched SNPs. We contrast findings with 11 genome-wide significant variants for tobacco smoking. To document generalizability, we generate and test a random phenotype.

**Results:** None of the 5 ML algorithms predict OUD better than chance when ancestry was balanced but were confounded with ancestry in an out-of-sample test. In addition, the algorithms preferentially predicted admixed subpopulations. Random sets of variants matched to the candidate SNPs by allele frequency produced similar bias. Genome-wide significant tobacco smoking variants were also confounded by ancestry. Finally, random SNPs predicting a random simulated phenotype show that the bias attributable to ancestral confounding could impact any ML-based genetic prediction.

**Conclusions:** Researchers and clinicians are encouraged to be skeptical of claims of high prediction accuracy from ML-derived genetic algorithms for polygenic traits like addiction, particularly when using candidate variants.

## 1. Introduction

Machine learning (ML) applications are increasingly used to leverage big data from electronic health records to classify patient populations (Ellis et al., 2019). In the realm of direct to consumer (DTC) and physician-guided genetic testing, ML approaches are gathering momentum, especially for psychiatric disorders. Currently, several commercial entities offer genetic testing for psychiatric disorders, and some have begun to offer controversial and scientifically disproven proposals for genetic embryo selection for behavioral and psychiatric traits (Karavani et al., 2019). While most genetic tests within psychiatry are aimed at medication efficacy in patients (e.g., pharmacogenetic or pharmacokinetic testing), a few recent tests target prediction of future psychiatric disorders. Alongside the potential ethical challenges of such predictions (Hooker, 2021) lie the scientific limitations. The genetic “inputs” that are used by these tests typically comprise of “candidate gene variants” that are scored using pattern recognition software, powered with “artificial intelligence” frameworks, such as machine learning (ML). Most candidate variants have not borne out in unbiased genome-wide association studies (GWASs) (Border et al., 2019; Johnson et al., 2017). Yet, they tend to be popular as genetic markers of disease risk in commercial assays (e.g., Keri Donaldson, et al. 2017). Exacerbating the problem of false positive candidate variant findings, past work on ML algorithms in psychiatric genetics has shown that these models may not systematically account for genetic ancestry (Bracher-Smith et al., 2020). As a consequence, patients and physicians are now confronted with products that may be unrelated to disease, but rather serve as panels of ancestry informative markers.

One psychiatric illness that is being targeted by ML-based genetic algorithms is opioid use disorder (OUD), a complex trait associated with high disease burden, and estimated to affect 2% of the adult population (Saha et al., 2016). Predictive tools that aim to identify at-risk individuals for prevention and early intervention are being developed (Ellis et al., 2019), and because OUDs are moderately heritable (h^2^ = 30-70%; Sun et al., 2012), incorporating genetic variation into a predictive tool has great appeal. In addition, because opioids comprise front-line pain management drugs, biomarkers that index risk of OUD in this setting are of potential interest. Industry-selected candidate variants (e.g., in dopamine and serotonin candidate genes) are routinely favored by those developing purported prediction tools for addiction, despite the scientific consensus regarding the weaknesses inherent to selection of candidate genes (Border et al., 2019; Duncan and Keller, 2011; Johnson et al., 2017). However, OUD is highly polygenic with a large number of variants of small effect contributing to its heritability. The largest genome-wide association study (GWAS) of OUD to date (15,756 OUD cases and 99,039 controls) identified only one genome-wide significant variant, rs1799971, in the gene encoding the mu opioid receptor (*OPRM1*) (Zhou et al., 2020); the effect size associated with this variant was small (β = -0.066 [SE = 0.012]). Current estimates of the total single nucleotide polymorphism (SNP)-based heritability of OUD is 11% (SE = 1.8%)(Zhou et al., 2020), putting a limit on overall predictive ability using common variants. Thus, we hypothesized that when ML algorithms utilize unsubstantiated candidate variants and do not properly account for population stratification, they produce predictions of disease outcome that are spurious and can cause mis-diagnosis.

In contrast to OUD, loci for tobacco phenotypes are numerous (Liu et al., 2019). Similar to OUD, the top genome-wide significant variants for tobacco smoking were in candidate genes (e.g., variants in *CHRNA5, CYP2A6*). These GWAS-validated candidate variants may have measurable impact on smoking cessation (Chen et al., 2020). Whether the selection of GWAS-validated candidate variants ameliorates challenges of ancestrally confounded ML prediction remains unevaluated. To investigate whether genome-wide significant variants that may also have been candidate variants would overcome limitations of the OUD genetic test, we selected the 11 lead SNPs from the largest GWAS of cigarettes smoked per day for a comparison analysis (Liu et al., 2019). To further demonstrate the generalizability of ancestral confounding in ML, we simulated random genotypes and phenotypes to document that ancestral confounding produces seemingly accurate prediction even when the phenotype is random noise and the variants are selected randomly from the genome.

OUD and tobacco smoking are leading contributors to mortality – precision medicine efforts to preempt progression to drug misuse or intervene with tailored treatments will likely continue to incorporate genetic data. While a good predictor could aid physicians by providing them with additional information on which to base personalized treatment options, inaccurate predictive tests pose substantial hazards. For OUD in particular, the possible harms attributable to a false positive result include both the withholding of beneficial medication and discrimination (e.g., employer bias). Such tests must be rigorously evaluated. Here, we examine two critical considerations in genetic prediction tools, particularly those developed using ML: population stratification and variant (feature) selection. First, we test the prediction accuracy of ML models with candidate variants that comprise a commercial genetic test (previously Life Kit Predict from Prescient Medicine, now available from Solvd Health: https://solvdhealth.com/oud/). We examine accuracy as genetic ancestry is progressively accounted for. We also test the impact of mismatched ancestry as a predictor of degree of admixture in African Americans. Next, to test if a better choice of genetic variants ameliorates the problem, we select 11 variants implicated by the largest GWAS of cigarettes per day and use the same ML models to predict tobacco dependence, as assessed with the Fagerström Test for Nicotine Dependence. We also use random sets of genetic variants (that have been matched by allele frequency to the original variants) to see how these randomly selected SNPs compare to the candidate SNPs across different subsamples for a simulated trait. We hypothesize that ML algorithms will preferentially identify ancestral confounding over traits of interest, regardless of the quality of the genetic variants.

## 2. Methods

### 2.1 Selection of direct-to-consumer test methodology for comparison

We identified current consumer/physician-oriented genetic tests by conducting a web search within Google for “Genetic Testing for Psychiatry” and “Genetic Testing for Addiction” and selecting all tests from the first five pages. **Supplemental Table 1** shows a list of the 12 tests that were found, and known mechanisms for evaluation. The methodology presented in the current study was based on those used by Solvd Health (previously Life Kit Predict®), a physician guided genetic testing kit for OUD. This test was selected because (a) it purports to predict, with 97% accuracy, risk for OUD, a complex trait that has been shown to be highly polygenic and influenced by environment, (b) is accompanied by a training procedure that was published and therefore, can be recapitulated. While not completely transparent, this was the only test with enough information to allow us to evaluate the test, as described below.

The genetic component of Solvd Health’s prediction algorithm relies on 15 or 16 candidate single nucleotide polymorphisms (SNPs) depending on the version of the test (Donaldson, et al. 2017), most of which are used in other DTC tests (see **Supplemental Table 1**) and have often been labeled in the psychiatric genetics literature as “candidate genes” (Border et al., 2019; Johnson et al., 2017). Only one of these SNPs (*OPRM1*\*rs1799971) has been shown by GWAS to affect OUD risk; the very small effect size of this variant (beta= -0.066, p=1.51e-10) is unlikely to predict OUD risk to a great degree. With the exception of rs1799971, none of these candidate SNPs have been associated at genome-wide significant levels (p<5E-8) (**Supplemental Table 2**) with *any* complex trait in the GWAS Atlas (Watanabe et al., 2019) (**Supplemental Table 3**). However, the MAFs of many of the candidate SNPs vary greatly across ancestral populations (**Figure 1**). That is, taken individually, they tend to be associated with one’s *ancestral population*, but not to a *trait*. Accordingly, it was our expectation that sets of these markers would also necessarily be associated to population rather than trait, regardless of the sophistication of the interposed statistical methodology.

**Figure 1.**
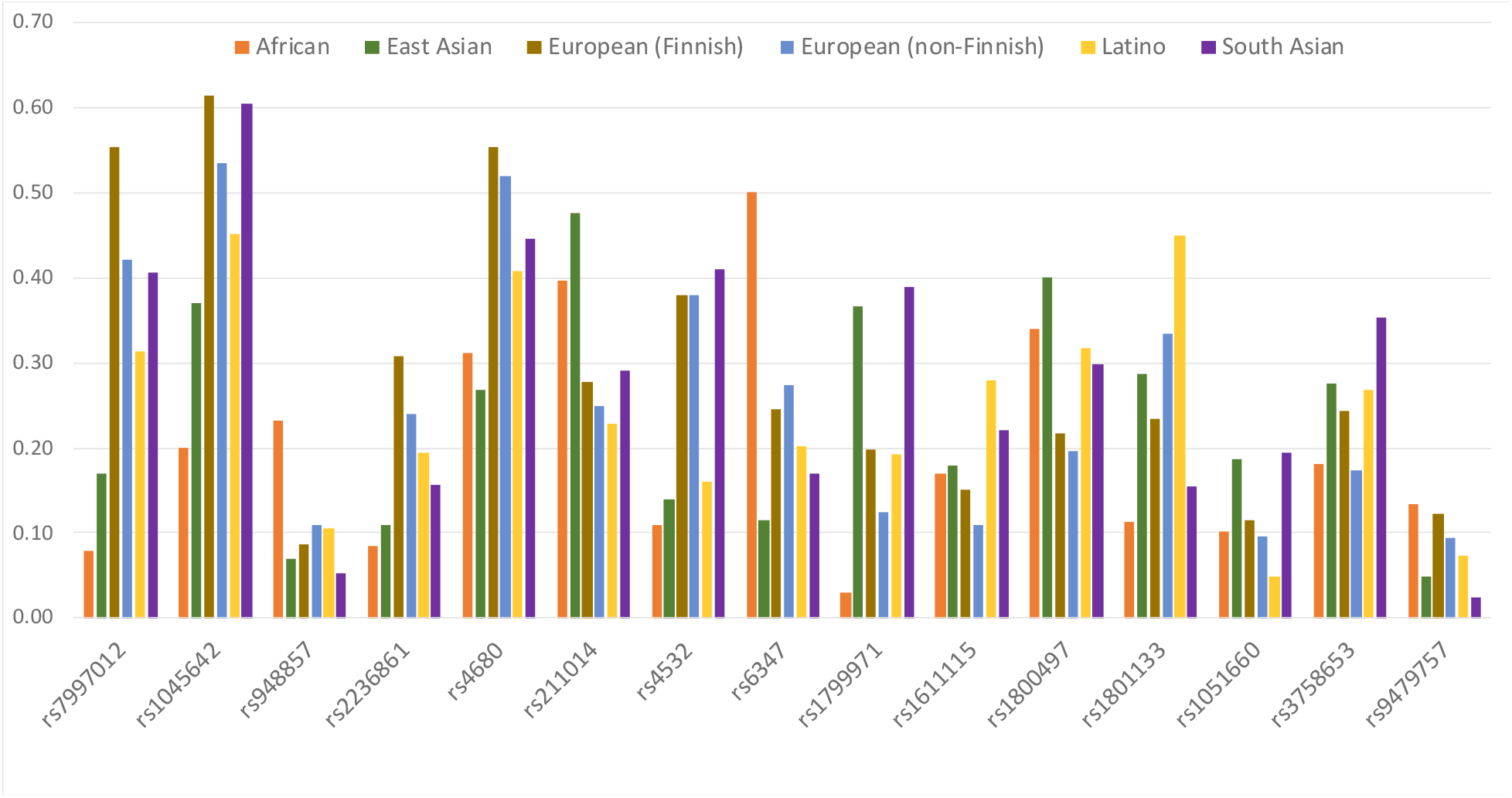
Population allele frequencies (from GNomad(Karczewski et al., 2020)) for the candidate alleles in Donaldson et al.(Keri Donaldson, Laurence Demers, Joe Lopez, 2017) LifeKit Predict® (https://prescientmedicine.com/technologies/lifekit-predict/; accessed September 8th, 2020) across different major geographic ethnic groups showing substantial variation in frequency across global populations.

### 2.2 Sample description

We tested the approach using data from subjects recruited at five sites across the eastern United States as part of the Yale-Penn study of the genetics of substance dependence and comorbid psychiatric and behavioral phenotypes (McCarthy et al., 2016). All participants were interviewed with the Semi-Structured Assessment for Drug Dependence and Alcoholism (Consortium et al., 2016) and provided written informed consent through a protocol approved by the institutional review board at each participating site – Yale Human Research Protection Program (protocols 9809010515, 0102012183, and 9010005841), University of Pennsylvania Institutional Review Board, University of Connecticut Health Center Institutional Review Board, Medical University of South Carolina Institutional Review Board for Human Research, and the McLean Hospital Institutional Review Board. Funding agencies did not play any role in the study.

### 2.3 Genotyping and quality control

The Yale-Penn phase 1 sample was genotyped using the Illumina HumanOmni1-Quad array. Individuals with mismatched sex or genotype call rate < 98% were removed; SNPs with genotype call rate < 98% or minor allele frequency < 0.01 were removed before imputation. Imputation was performed using Minimac3 (McCarthy et al., 2016) and the Haplotype Reference Consortium reference panel implemented in the Michigan Imputation Server (https://imputationserver.sph.umich.edu/index.html). More details on the Yale-Penn sample can be found elsewhere (Zhou et al., 2017).

Genetic ancestral group was defined by principal component (PC) analysis on genotyped SNPs (pruning by linkage disequilibrium of r^2^ > 0.2) and the 1000 Genome phase 3 reference panels (Auton et al., 2015) using EIGENSOFT (Patterson et al., 2006; Price et al., 2006a). The first 10 PCs were used to cluster the participants into African-American and European-American groups and to remove outliers from the 2 groups.

European-ancestry proportions in African-American samples were estimated using ADMIXTURE (Alexander et al., 2009). SNPs were included in ancestry prediction following the developer’s recommended independent SNP selection procedures. ADMIXTURE’s cross-validation (CV) procedure was used to determine the most appropriate K – the most sensible number of component ancestries with which to model unknown sample ancestries. Based on lowest CV error and failure to substantially reduce CV error with additional K, we chose K=2 as appropriate for these data. The mean European ancestry for the African-Americans included in this study was 23.2%.

### 2.4 Selection of random sets of 16 variants

We calculated, in the Yale-Penn sample, the minor allele frequency (MAF) of the SNPs in Donaldson et al. (2017) using PLINKv1.90 over a total sample of 5,057 individuals (3286 African American and 1768 European American). For comparison, in addition to the SNPs used by Donaldson et al. (2017) we identified 8 lists of random SNPs -each SNP from Donaldson et al. was replaced by a random SNP with matched MAF in the 2 populations; all random SNPs were unique.

### 2.5 Machine learning training procedure

Each of the 16 alleles was dummy coded for homozygosity or heterozygosity status to allow for interactions among different levels of dosage of each minor allele from each SNP with minor allele dosages of other SNPs (i.e., to measure epistasis, if present). Each supervised ML algorithm was trained separately in the same training set, which varied from 500 to 1000 individuals based on k iterations of the learning curve (see **Figure 2**). The training and test sets were initially analyzed in a way that assured that they were completely confounded by population differences, with all cases of European ancestry and all controls of African ancestry. At each iteration of the learning curve, we added 10 individuals of African descent to the cases and 10 individuals of European descent to the controls, through 26 steps, to reach completely balanced samples (see **Figure 2**).

**Figure 2.**
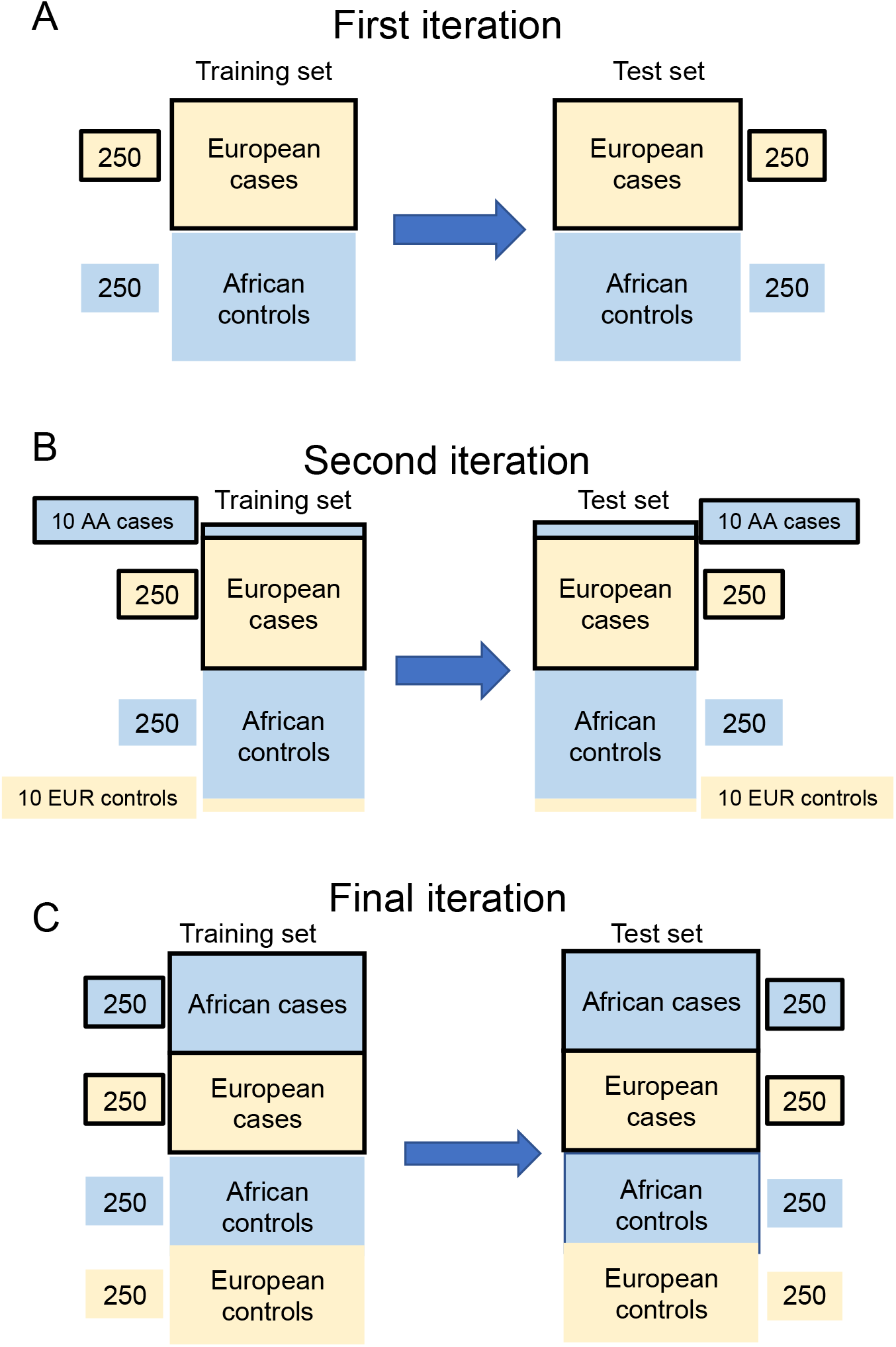
Learning curve procedure for all analyses. We had a complete and non-overlapping training and test set, each of 1000 subjects with 250 cases and controls of European and African descent. (A) For the first iteration we started with 250 subjects of European descent (tan) that were OUD cases, and 250 subjects of African descent (blue) that were controls. (B) At each iteration, we added 10 OUD individuals of African descent to the cases and 10 controls of European descent to the controls. We estimated the model in the training data and used it to predict OUD status in the non-overlapping test set. (C) By the final iteration, we had a training and test set that was balanced by major geographic ancestry and OUD status.

For our analysis, we chose 5 broad algorithm-generating methods as a survey of supervised ML, because they have been used by academic and commercial entities to attempt to predict OUD. All were implemented in the Caret package in R version 3.6.110: (1) (extreme) Gradient Boosted Machines (GBM), which incorporate population stochastic gradient descent procedures that are ubiquitous in industry; (2) Linear and (3) nonlinear (radial basis function) Support Vector Machines (SVM) to compare predictive accuracy from different kernels; (4) Random Forests (RF) to represent more complex tree structures; and (5) Elastic Nets (EN) for representation of (flexible) linear regression models. All models were trained with 10-fold cross validation and a hyperparameter grid-search in the training set. All Area Under the Curve (AUC) and pseudo r^2^ were extracted from the non-overlapping test set. Learning curves were plotted at all iterations.

### 2.6 Generalizability of confounding tested using Tobacco Dependence and a random binary phenotype

To examine the generalizability of this confounding beyond the test case of OUD prediction, we examined prediction of tobacco dependence, measured by the Fagerstrom Test of Nicotine Dependence (FTND). Cases were defined as scores ≥4 and controls with scores <4 (Heatherton, et al., 1991). Eleven genome-wide significant lead variants for cigarettes per day were selected from the Liu et al. (2019) cigarettes per day GWAS and the Quach et al. (2020) GWAS of FTND. We used both the cigarettes per day and the FTND GWAS because they are genetically correlated at .95 and share some of the same lead SNPs (Quach et al., 2020): rs3743078, rs16969968, rs56113850, rs58379124, rs215600, rs3025383, rs2072659, rs7951365, rs7431710, rs11725618, rs45568238. We limited our learning curves to 140 subjects in each ancestral group and case/control group (instead of 250), as this sample size allowed for ancestral balance needed for the learning curve approach (as there are only 140 control of European ancestry available). We also trained the models within the European American (EA) and African American (AA) subsamples. Data on 658 cases (AA = 302, EA = 356) and 342 controls (AA = 198, EA = 142) were used in training and testing sets, respectively.

To explore the problem further, we generated a random binary phenotype by drawing from a binomial distribution. That is, the phenotype was essentially random noise and therefore not truly predictable. We matched the number of cases and controls for our random variable by genetic ancestry, such that we ended with the same split in cases and controls by ancestry as was used in our OUD demonstration. We then took the 8 random SNP set permutations and used them to predict the random noise. Because we used random SNPs with random outcomes, the effects provide an empirical NULL hypothesis: what the data look like when the result is by definition meaningless. We hypothesized that this empirical null will still show high effect sizes at high confounding, i.e., even random noise can seemingly be “accurately” predicted when the sample is confounded.

## 3. Results

### 3.1 Evaluation of a modern OUD prediction kit in the presence of confounding by ancestry

For our empirical test, all models were trained using the panel of 16 SNPs referenced in Donaldson et al. (Donaldson, et al. 2017), the basis for Solvd Health’s OUD test kit, and were trained to predict OUD in the Yale-Penn sample. These 16 variants demonstrate substantial allele frequency differences across ancestries (**Figure 1)**. As shown in **Figure 3A** for all 5 ML methods, prediction of OUD case status was apparently high (AUC > 0.8) when the sample was fully confounded (that is, when predictions were essentially predictions of genetic ancestry), and case-status prediction decreased as samples were better ancestrally balanced, until the prediction was no better than expected by chance alone in a balanced sample (AUC approached 0.5). At every iteration of every ML approach, the 16 variants predicted genomic ancestry much better than they predicted OUD.

**Figure 3.**
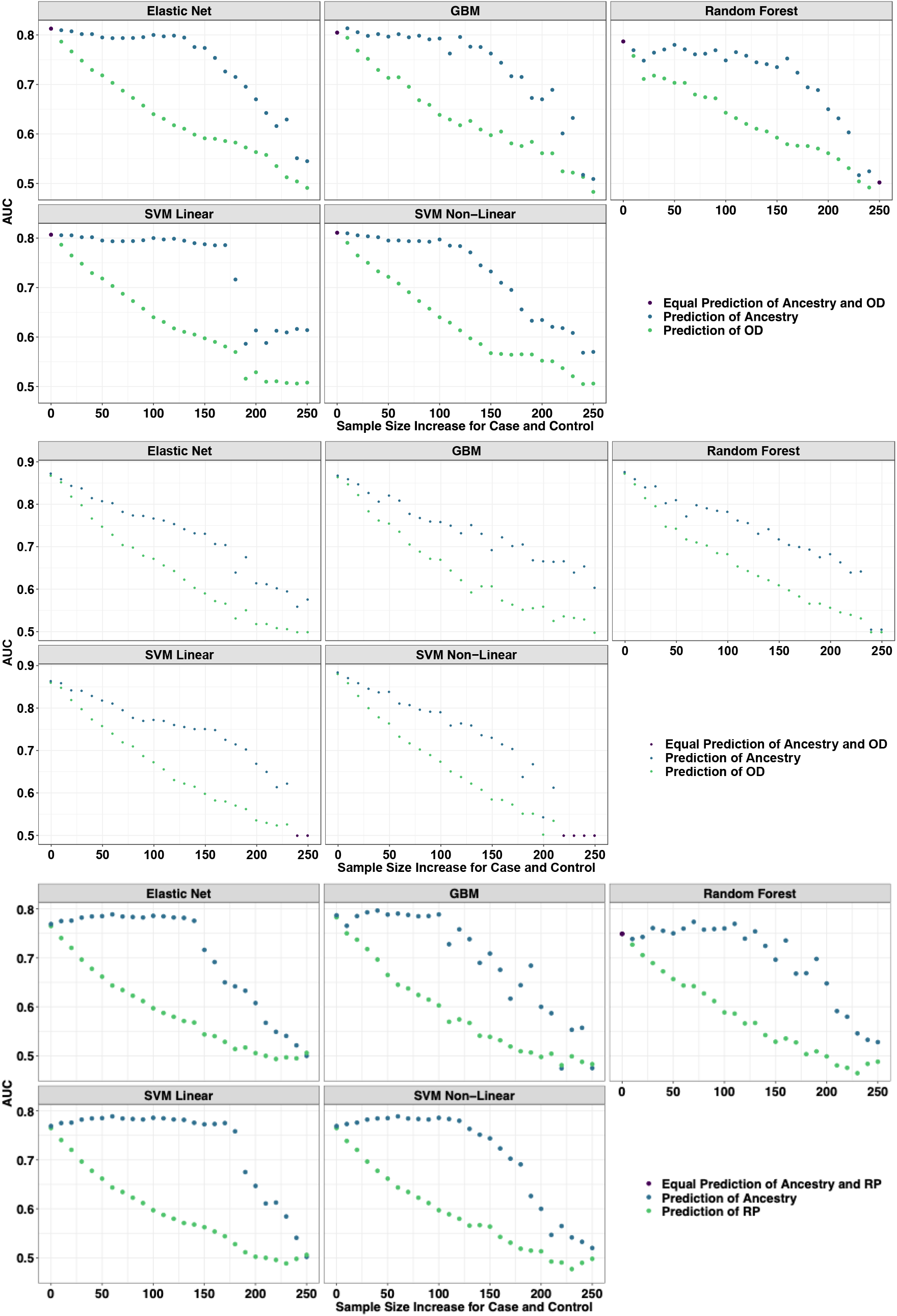
Learning curves from models trained to predict opioid dependence from 16 “reward-related” SNPs (Donaldson et al., 2017). The curves are plotted by AUC based on their prediction of opioid dependence (orange) and geographic ancestry (blue) as the samples start from complete population confounding become more balanced by major geographic ancestry (European American or African American) until completely balanced. Each data point represents a larger and more balanced sample size by adding 20 individuals, 10 African American cases and 10 European American controls (as measured on the x-axis). **(A)** *A priori* Candidate SNPs predicting Opioid Use Disorder. **(B)** Set 1 of randomly selected (MAF matched) SNPs predicting Opioid Use Disorder. **(C)** Set 1 of Random (MAF match) SNPs predicting a random phenotype binary phenotype. Across all perspectives, the prediction is entirely driven by major geographic ancestry.

### 3.2 Random SNPs predict OUD as well as biologically plausible SNPs due to confounding

All iterations with 8 permutations of random SNPs matched on minor allele frequency to those in Donaldson et al. (2017) performed similarly to the published candidate SNPs (**Figure 3B** shows 1 permutation, the other 7 are shown as **Supplemental Figure 1**). Across all iterations of all permutations the ML models using random SNPs were apparently predictive of OUD when confounded by ancestry, with decreasing prediction as ancestral balance improved. They were better predictors of ancestry than OUD. Therefore, the candidate variants perform no better than randomly selected variants with the same ancestral allele frequencies.

### 3.3 Confounded models are better at detecting subpopulation within minority populations than diagnosis

As African Americans include substantial European admixture (Jordan et al., 2019) we examined whether the 16 OUD variants used by Solvd Health predict the extent of European (genetic) admixture within the African American cases and controls. We chose the 15^th^ iteration (**Figure 3**) of the learning curve as it had the greatest balance of ancestry that still offered some prediction of OUD that was greater than chance. Across all approaches, ML models designed to predict OUD were up to 5 times better predictors of the percent of European admixture in African-American individuals than of OUD (i.e., case status) (**Figure 4**).

**Figure 4.**
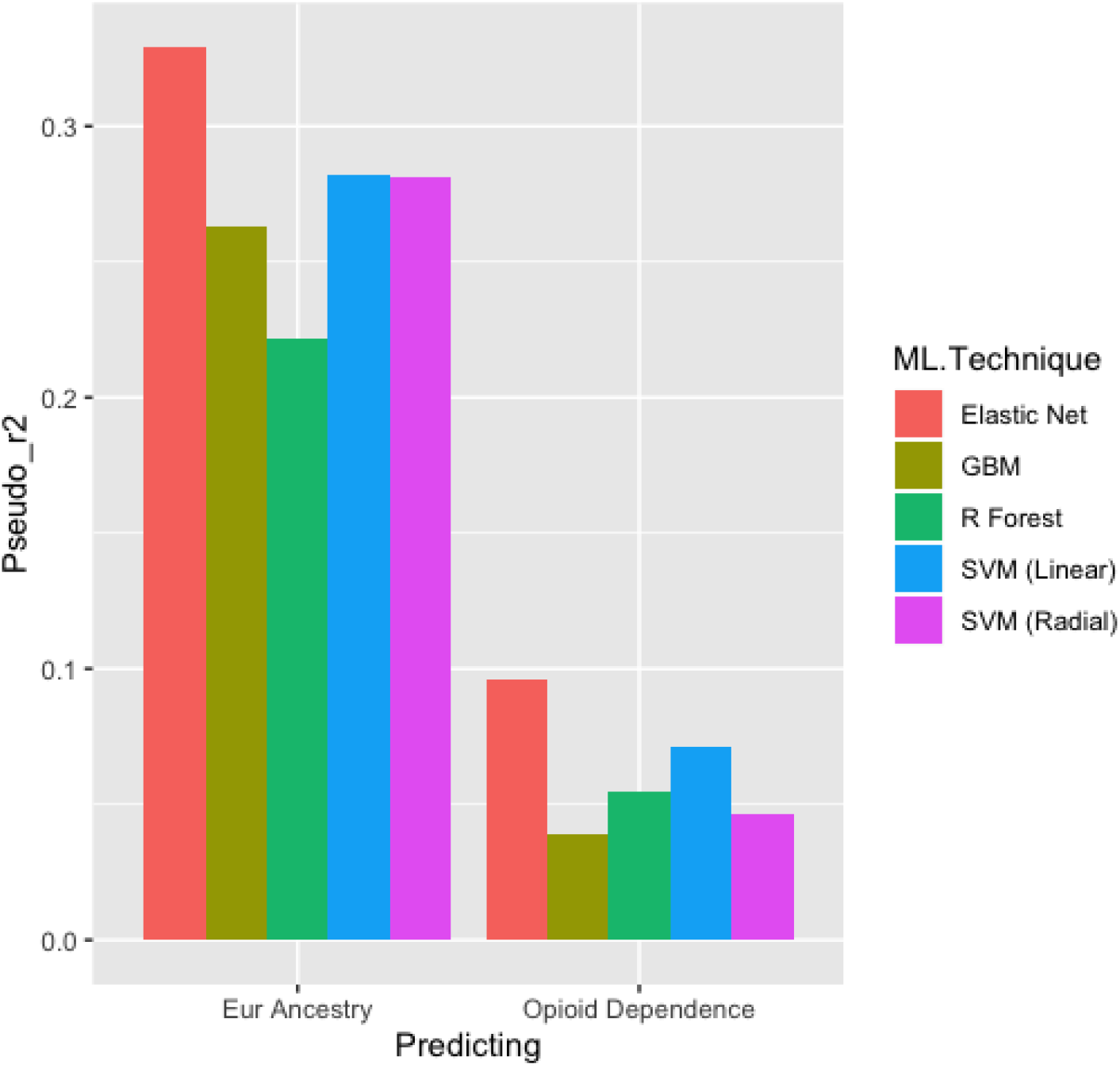
Bar plots of the pseudo r^2^ from a logistic regression comparing the predictions of opioid dependence and percentage of European ancestry in a sample of 250 African American individuals from the Yale-Penn Test set. Pseudo r^2^ was used instead of AUC because the percentage of European descent is a continuous variable and this put both predictions on the same scale.

### 3.4 Genome-wide significant variants for Nicotine Dependence show similar bias

Using variants discovered in a large, un-biased GWAS of tobacco smoking (Liu et al., 2019; Quach et al., 2020) showed the same pattern: when models were confounded by ancestry, genetic prediction of nicotine dependence appeared to be accurate but the apparent accuracy fell to zero as we balanced ancestral groups within cases and controls **(Supplemental Figure 2)**. At all iterations, the model was better at predicting ancestry than nicotine dependence, even with SNPs that were genome-wide significant. Finally, we trained the prediction model within each ancestral population and found that AUC values did not exceed .504 in the European sample **(Supplemental Table 4**), suggesting no prediction even when training within the discovery ancestral population.

### 3.5 Ancestral confounding leads to random genotypes making apparently accurate predictions of random phenotypes, mirroring results for OUD

We next took the 8 random sets of SNPs (above) and used them to predict a randomly generated phenotype. For a randomly generated phenotype, at perfect confounding by major geographic ancestral group, we get high apparent predictive accuracy of random noise, but as we balanced the training and test sets by ancestry, our model performs as expected, with prediction no better than a coin flip **(Figure 3C)**. Across all iterations (**Supplemental Figure 3**), all models trained to predict random noise were stronger predictors of ancestry than random noise, suggesting that even if the outcome is meaningless, we can gain the appearance of meaningful results in the presence of ancestral confounding.

## 4. Discussion

ML models trained either on a handful of selected candidate variants or across the whole genome are strongly sensitive to confounding by genetic ancestry (Polimanti et al., 2015). We demonstrate these underlying problems for a specific genetic test for OUD, but our simulations demonstrate that the confound is generalizable – once ancestry is accounted for, these ML models offer no evidence of predictive ability greater than chance. Our findings argue for great caution when evaluating results of other ML-based genomic analyses that do not explicitly and fully account for ancestral confounding.

In the field of ML, our results fall under the study of “algorithmic bias” (Hooker, 2021). Examples in healthcare research outside genetics (Obermeyer et al., 2019) show that pattern recognition with little understanding of underlying phenomena (e.g., social population stratification) may produce biased results. Here, we demonstrated that in the context of genomic data, this algorithmic bias was generated by population stratification, a well-characterized phenomenon in statistical genetics (Price et al., 2006b) that is yet to be widely dealt with by ML in psychiatric genetics (Bracher-Smith et al., 2020).

As human genetics has shown, this challenge is surmountable. Most ML algorithms allow for some form of de-confounding, typically as a multi-step or multi-model procedures. While typical ML pipelines employ multiple algorithms and simply select the best, more extensive individual attention to the choice of algorithm is needed to evaluate confounding in the face of known covariates. Several avenues may be pursued based on the choice of a model. For example, in this paper (and in those models used by industry professionals) gradient boosted machines were used, which can (but have yet to) include sample weights in the model training procedure. Corrections for support vector machines also exist, and remove statistical dependence in the model training procedure (Li et al., 2011). Extensive work with each algorithm will best determine future routes for de-confounding, and needs to be an essential part of model training beyond just predictive accuracy. To cut across algorithms here, we show stratified analysis with learning curves. In particular, learning curves should be purposefully developed with ancestral stratification in mind to ensure that lingering cryptic admixture does not confound predictions. Finally, statistical procedures will only take us so far, and careful considerations of samples and confounding, as is standard in GWAS literature, is critical to reduce confounding in genetic testing practices. However, restricting analyses to one continental ancestry (e.g., Europeans only) is not the solution to ancestrally confounded analyses. While that may attenuate gross confounding, we show that cryptic admixture remains an issue. Instead, larger training and testing samples of diverse ancestral populations are needed to accelerate genomic discovery and ensure that when aggregated effect sizes are large enough, precision medicine will benefit all global communities (Martin et al., 2017).

Even with appropriate adjustment for admixture, it is unlikely that candidate variants that are not substantiated by well-powered GWAS will produce any meaningful prediction of OUD. Recent meta-analyses of depression (Border et al., 2019), schizophrenia (Johnson et al., 2017), and executive function (Hatoum et al., 2019) overwhelmingly show that the vast majority of candidate variants in psychiatry do not rise to levels of genome-wide significance. An exception is addiction, where GWAS have recapitulated candidate gene findings. Unfortunately, as shown by our analyses of nicotine dependence, even these genome-wide significant variants fail to circumvent the issue of ancestral confounding within ML models. An alternative might involve incorporating information across the genome. For instance, polygenic risk scores (PRS) for tobacco smoking have produced promising findings, including in clinical settings (Chen et al., 2018). However, even PRS with appropriate control for ancestry in the largest samples to date offer limited clinical utility (Liu et al., 2019).

Several limitations are noteworthy. First, the original publication by Donaldson et al. (2017), did not provide detailed characteristics of the samples in which the algorithms were developed and tested, nor the specific procedures used. This is not atypical for “proprietary” commercial products, but made it challenging to fully approximate their analytic pipeline; hence we tested 5 different ML approaches. Standards for reporting such product development, such as the Transparent Reporting of a multivariable prediction model for Individual Prognosis Or Diagnosis (TRIPOD)(Collins et al., 2015), would allow researchers to better evaluate claims of genetic prediction, particularly using methods such as machine learning. Second, we focused on one existing product and did not evaluate all possible methods for genetic prediction, for example, polygenic risk scores (PRS). As noted above, PRS offer an opportunity to study aggregated genome-wide genetic susceptibility, but have their own caveats (e.g., additivity; see: Bogdan et al., 2018; Dudbridge, 2013; Martin et al., 2017). This work focuses squarely on one approach for genetic prediction as it increasingly gains momentum in allied health fields – use of candidate genes and ML. Limitations notwithstanding, this work shows that using a handful of candidate variants in a ML framework that is naïve to genetic confounders is likely to produce biased prediction that misclassifies individuals, especially in ancestrally mixed samples.

## 5. Conclusions

Opioids are useful for pain management, but are also highly addictive. Against the backdrop of the opioid epidemic, the desire for tests that can provide insight into the likelihood of patients developing OUD is understandable, and DTC or physician-guided testing seems appealing. However, to avoid problems of under-treatment, that might disproportionately affect people of admixed ancestry, it is critical that any proposed test be fully vetted to ensure that it properly accounts for potential confounding by ancestry and accurately predicts the trait of interest.

## Data Availability

Results from learning curves and ML scripts will be made available upon publication.

