## Supplemental Figure 1 for "Ancestry May Confound Genetic Machine Learning: Candidate-Gene Prediction of Opioid Use Disorder as an Example"

**Supplemental Figure S1.** We completed 8 permutations of MAF matched random SNPs for prediction of Opioid Dependence in the Yale-Penn dataset. The first permutation is **Figure 3** in the main text. Conclusions do not change based on the permutation, i.e. all analyses across all permutations are in-line with random SNPs performing as well as candidate SNPs and being highly confounded by ancestry.

**Permutation 2**

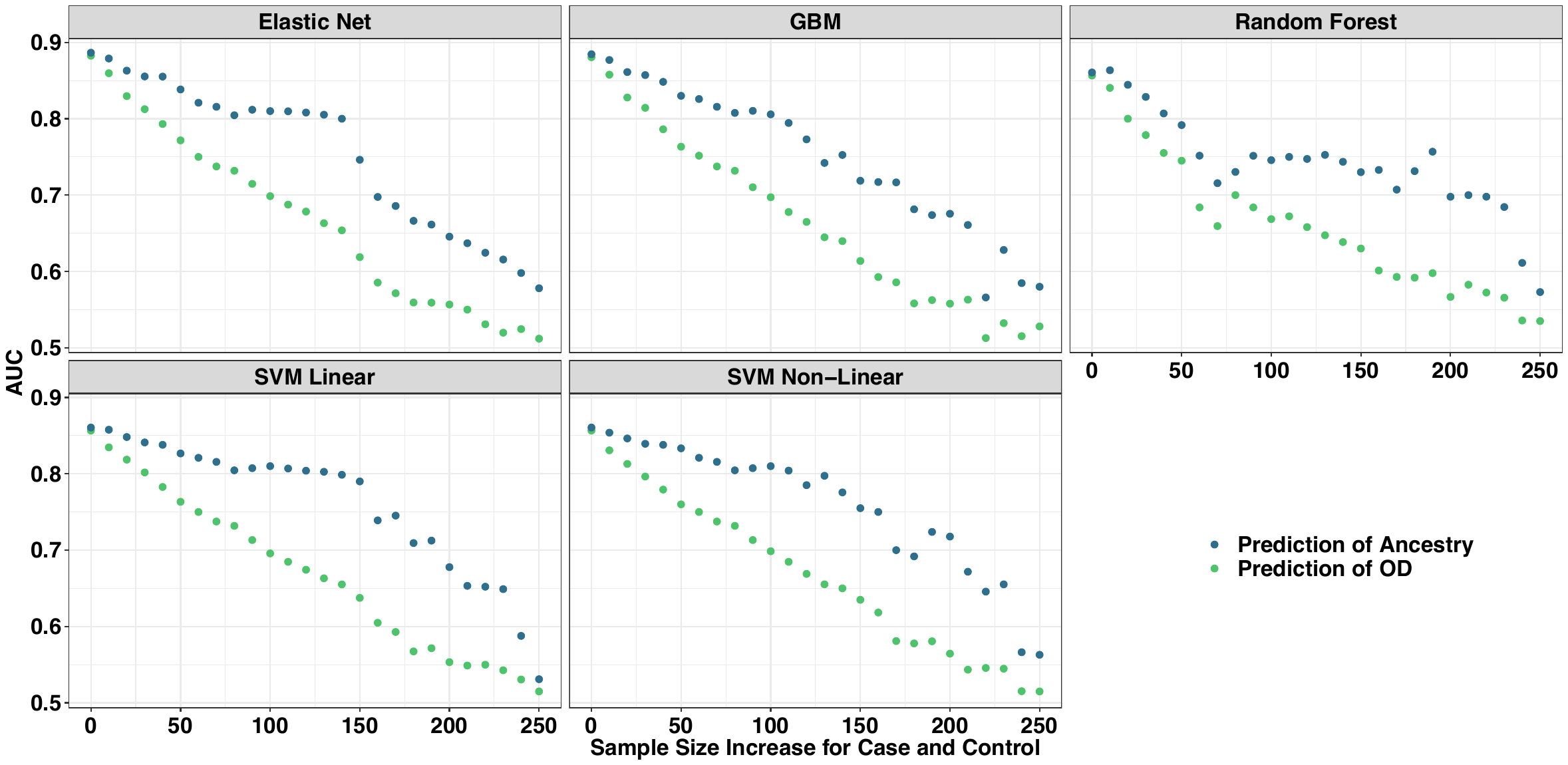

**Permutation 3**

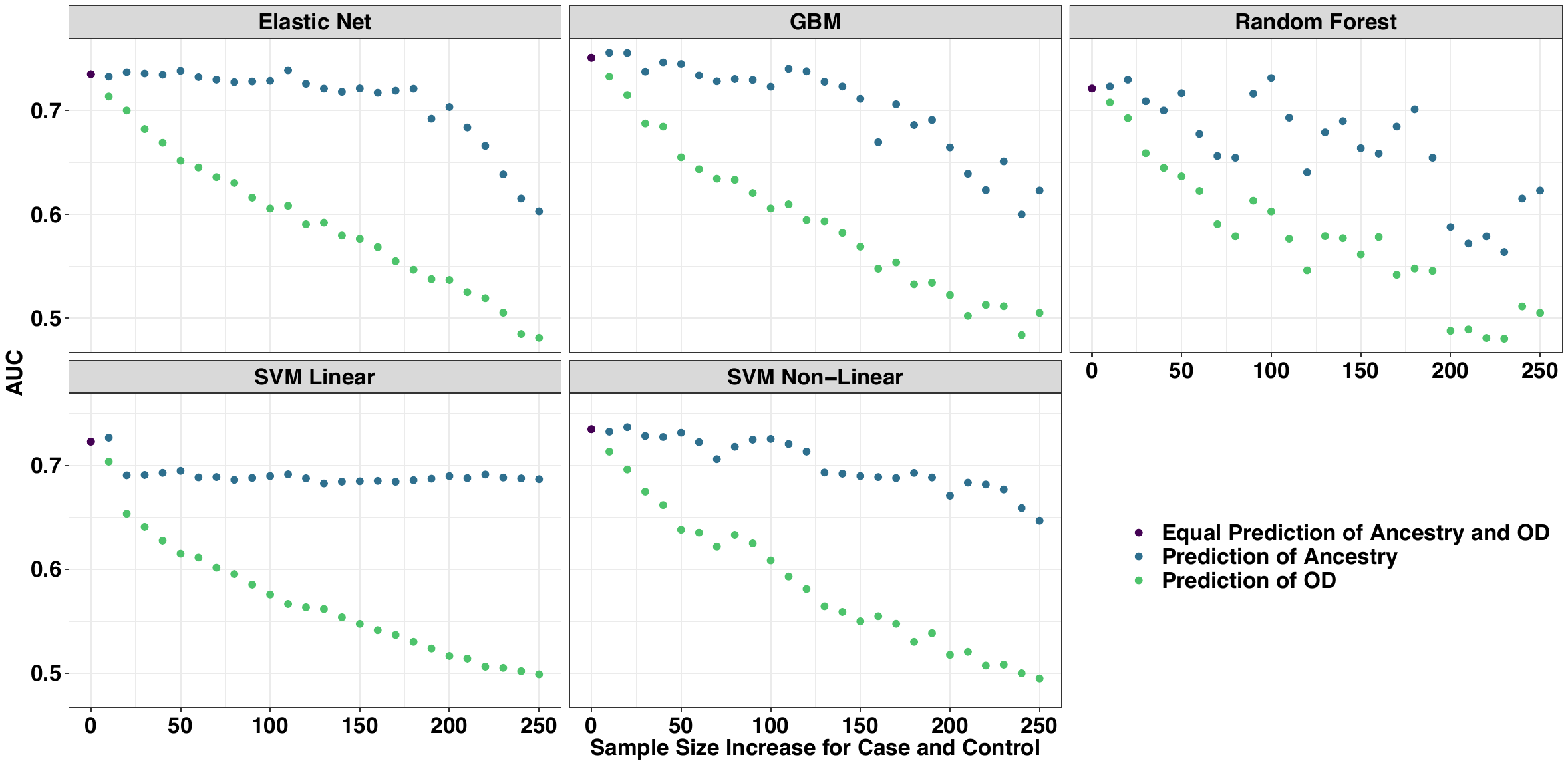

**Permutation 4**

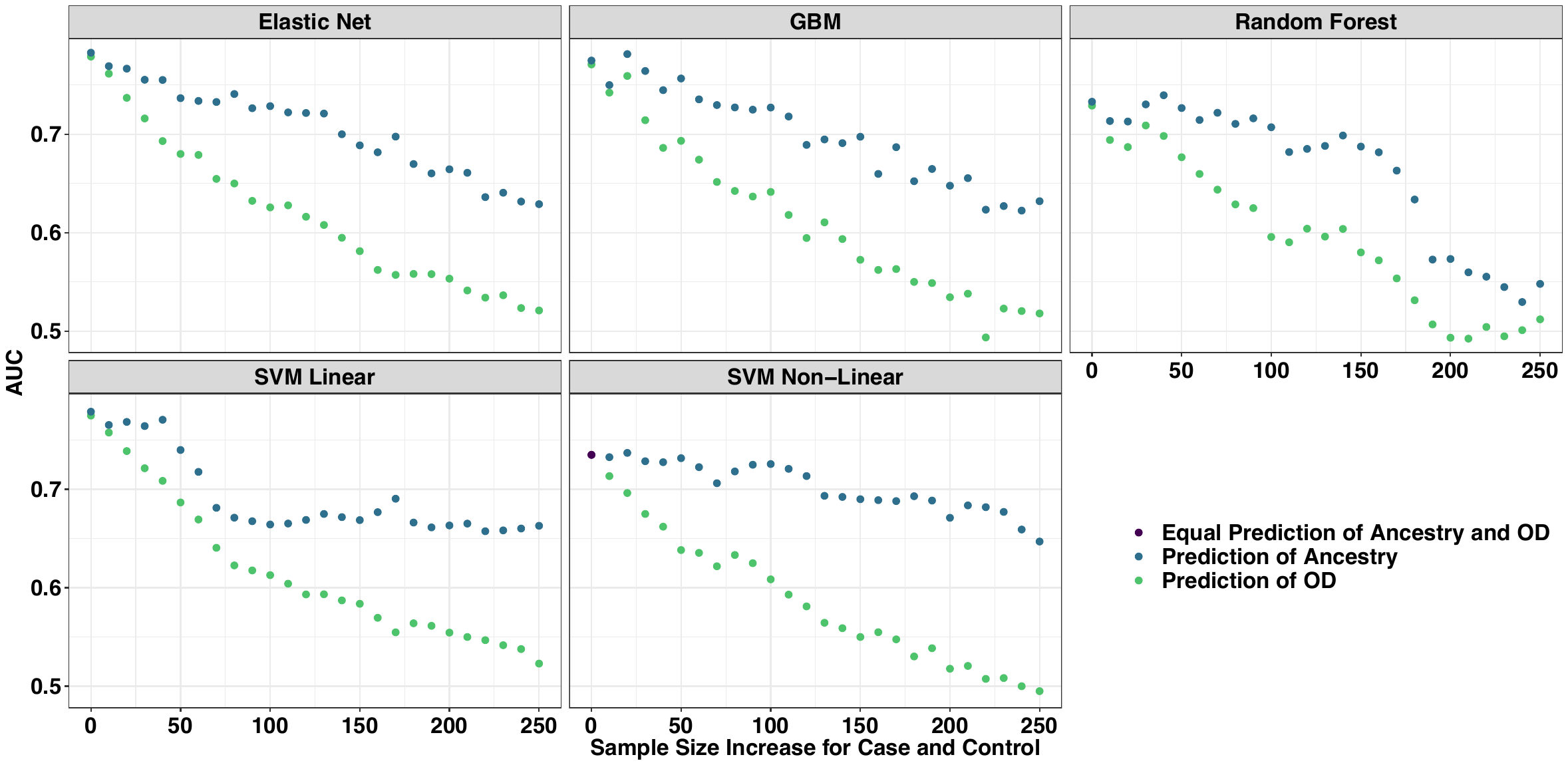

**Permutation 5**

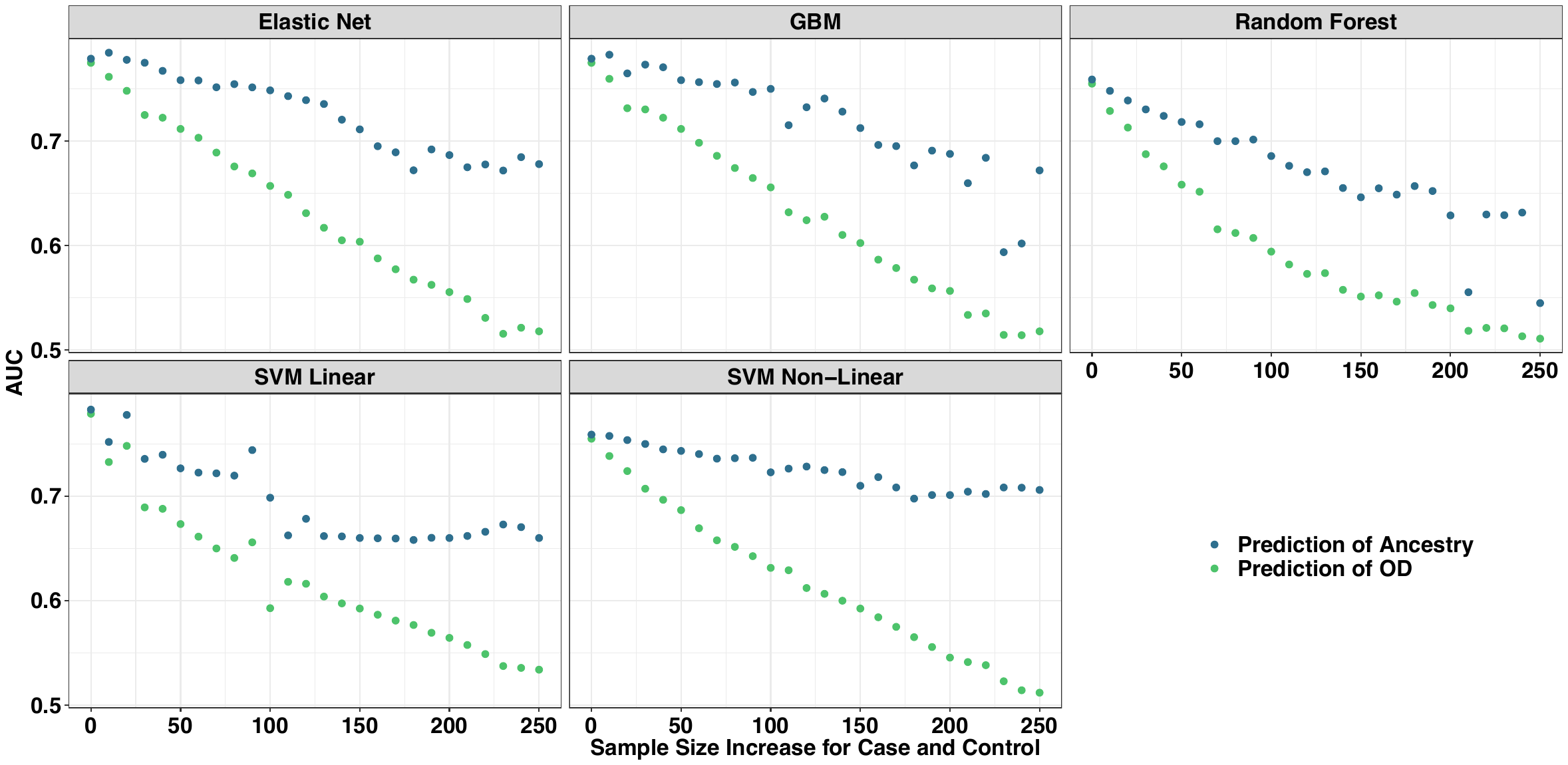

**Permutation 6**

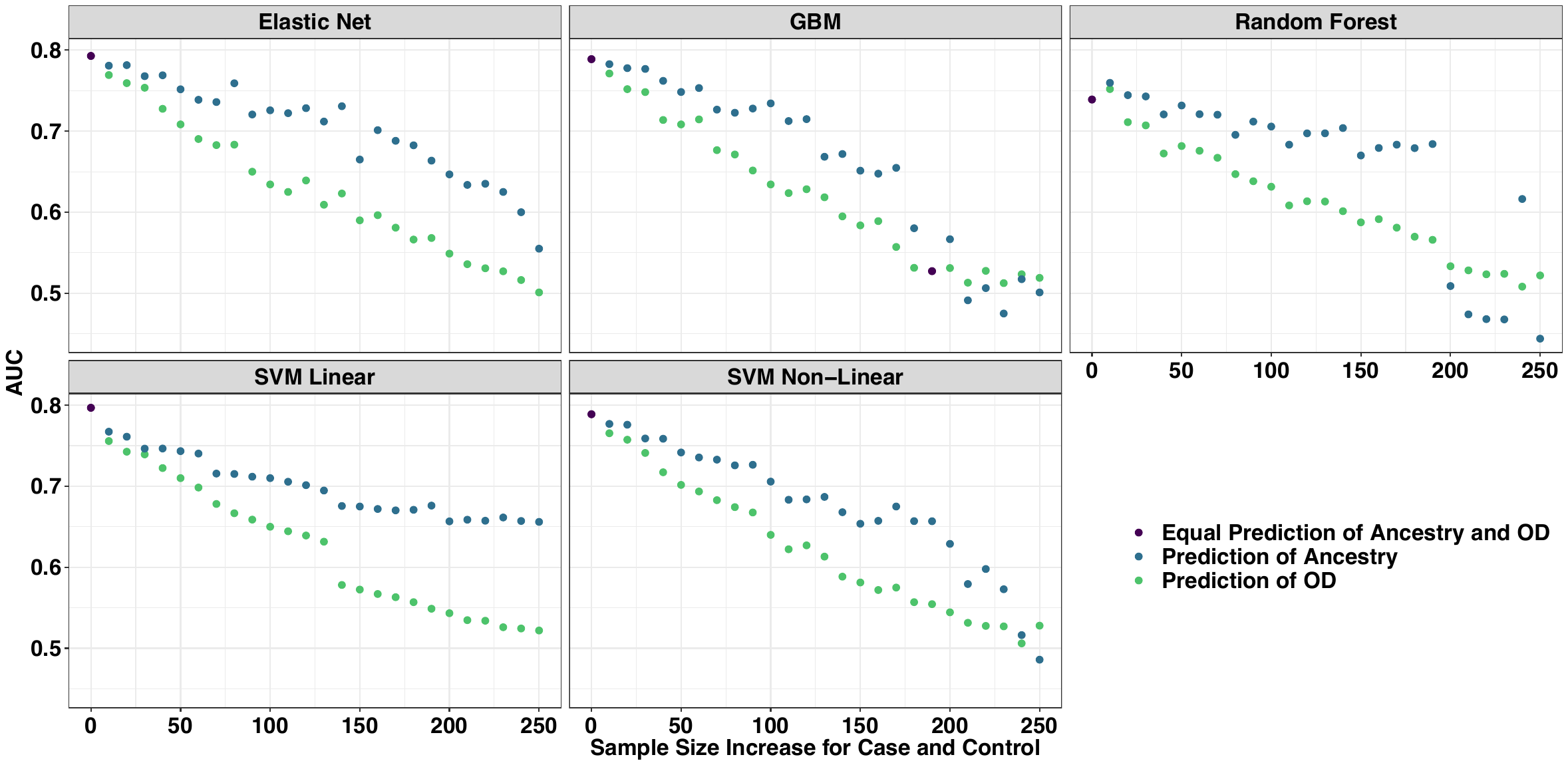

**Permutation 7**

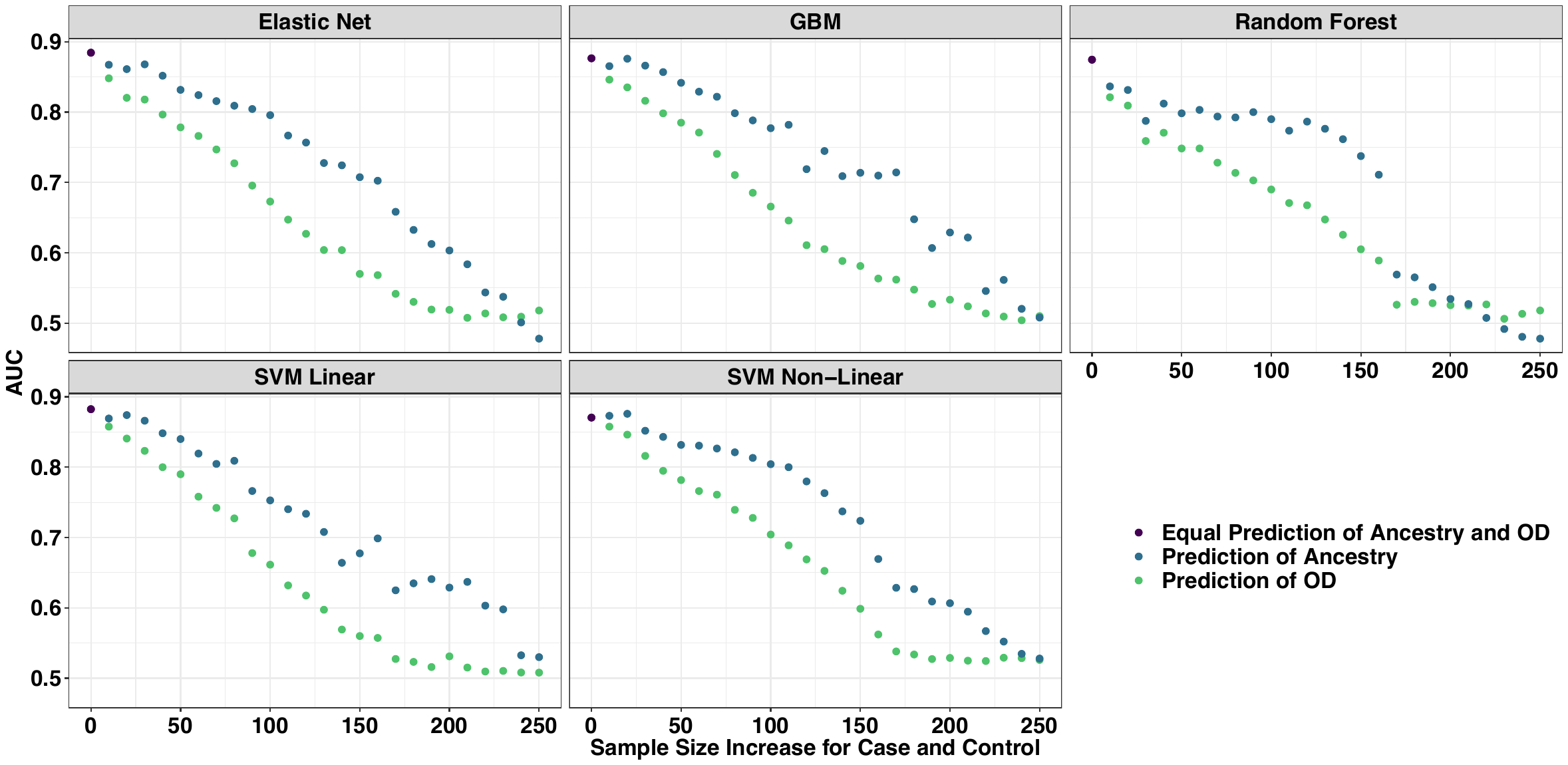

**Permutation 8**

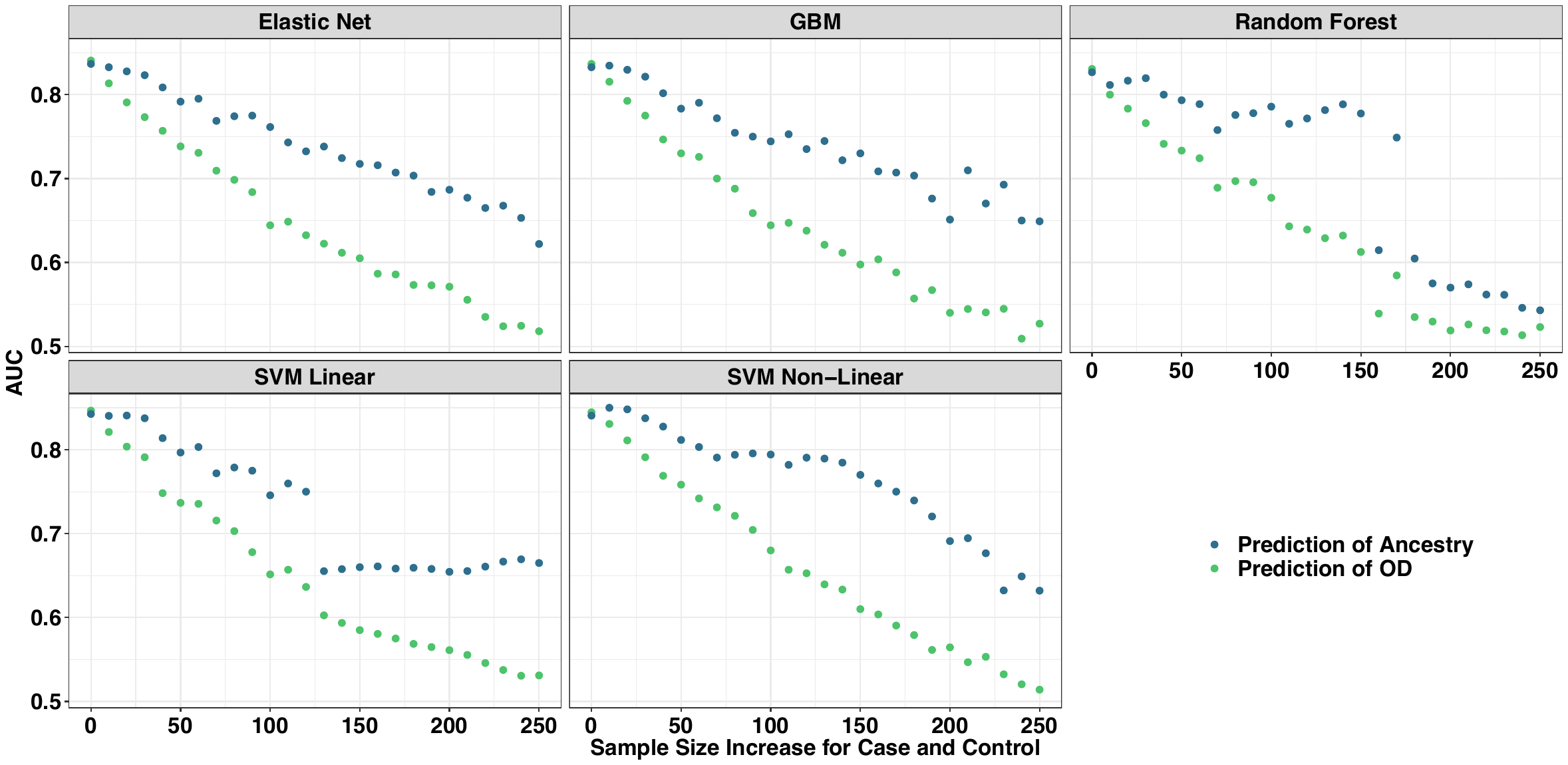
