## Supplemental Figure 2 for "Ancestry May Confound Genetic Machine Learning: Candidate-Gene Prediction of Opioid Use Disorder as an Example"

**Supplemental Figure S2.** We took 11 lead variants from GWAS of cigarettes per day and FTND GWAS and predicted diagnosis of tobacco dependence from the FTND using our ancestral informative learning curves. Even real SNP signals were highly confounded by ancestry, such as when the models were highly confounded they offered accurate prediction of tobacco dependence and became less effective as samples were balanced by ancestry.

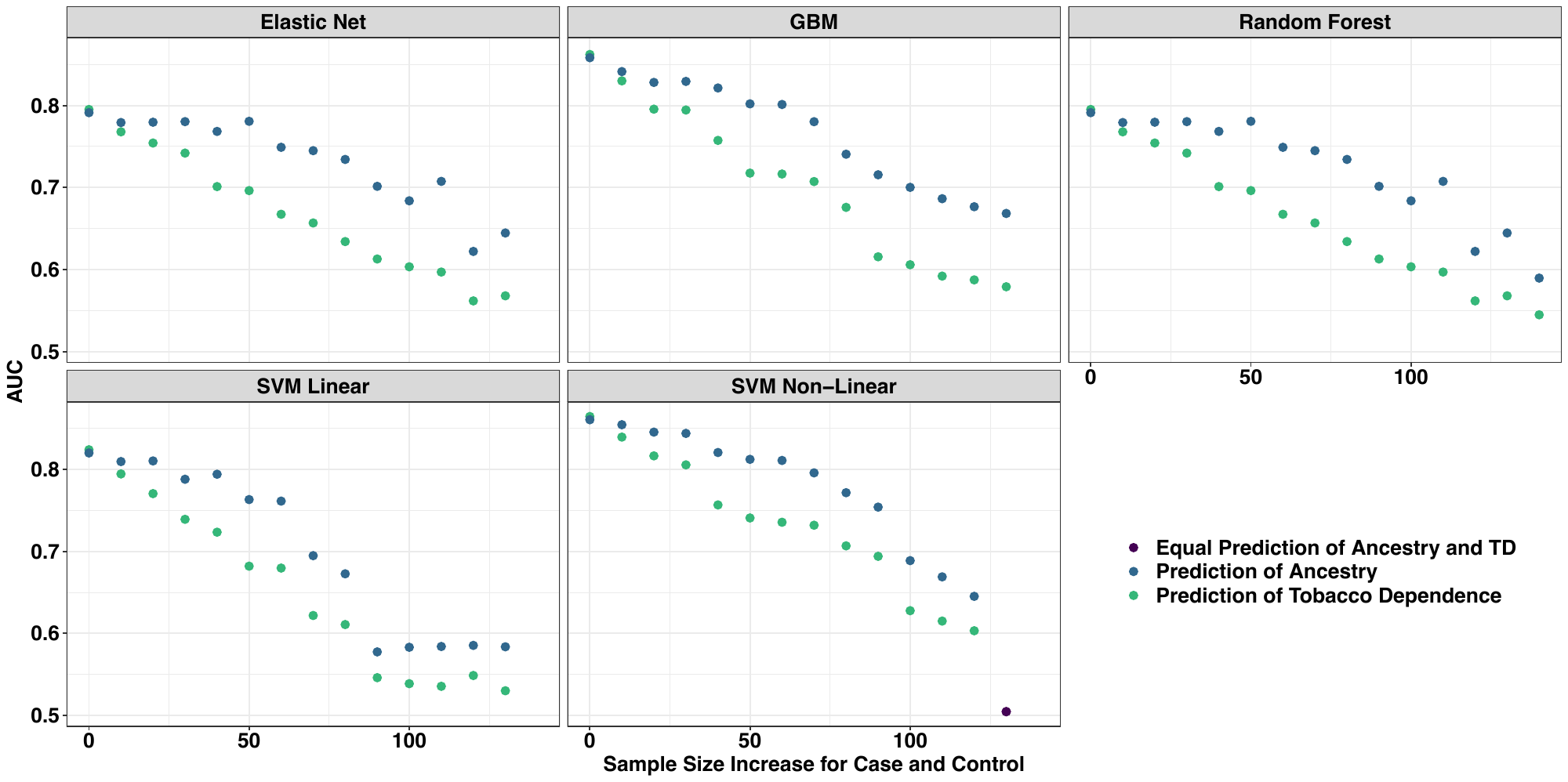
